# Safety of SARS-CoV-2 test-to-stay in daycare: a regression discontinuity in time analysis

**DOI:** 10.1101/2023.10.11.23296808

**Authors:** Felix Dewald, Gertrud Steger, Irina Fish, Ivonne Torre-Lage, Christina Hellriegel, Esther Milz, Anja Kolb-Bastigkeit, Eva Heger, Mira Fries, Michael Buess, Niklas Marizy, Barbara Michaelis, Isabelle Suárez, Gibran Horemheb Rubio Quintanares, Martin Pirkl, Annette Aigner, Max Oberste, Martin Hellmich, Anabelle Wong, Juan Camilo Orduz, Gerd Fätkenheuer, Jörg Dötsch, Annelene Kossow, Eva-Maria Moench, Gustav Quade, Udo Neumann, Rolf Kaiser, Madlen Schranz, Florian Klein

## Abstract

**Background and Objectives:** Test-to-stay concepts apply serial testing of children in daycare after exposure to SARS-CoV-2 without use of quarantine. This study aims to assess safety of a test-to-stay screening in daycare facilities.

**Methods:** 714 daycare facilities and approximately 50,000 children ≤6 years in Cologne, Germany participated in a SARS-CoV-2 Pool-PCR screening from March 2021 to April 2022. The screening initially comprised post-exposure quarantine and was adapted to a test-to-stay approach during its course. To assess safety of the test-to-stay approach, we explored potential changes in frequencies of infections among children following the adaptation to the test-to-stay approach by applying regression discontinuity in time (RDiT) analyses. To this end, PCR-test data were linked with routinely collected data on reported infections in children and analyzed using ordinary least squares regressions.

**Results:** 219,885 Pool-PCRs and 352,305 Single-PCRs were performed. 6,440 (2.93%) Pool-PCRs tested positive, and 17,208 infections in children were reported. We estimated that during a period of 30 weeks, the test-to-stay concept avoided between 7 and 20 days of quarantine per eligible daycare child. RDiT revealed a 26% reduction (Exp. Coef: 0.74, CI:0.52;1.06) in infection frequency among children and indicated no significant increase attributable to the test-to-stay approach. This result was not sensitive to adjustments for 7-day incidence, season, SARS-CoV-2 variant, and socioeconomic status.

**Conclusion:** Our analyses provide evidence that suggest safety of the test-to-stay approach compared to traditional quarantine measures. This approach offers a promising option to avoid use of quarantine after exposure to respiratory pathogens in daycare settings.

## Introduction

During the coronavirus disease 2019 (COVID-19) pandemic, daycare facilities were closed to mitigate infections.^1,2^ Systematic screenings for severe acute respiratory syndrome coronavirus type 2 (SARS-CoV-2) infections contributed to daycare re-openings, counteracting the profound impact of stay-at-home orders on the development and health of children.^1,3–6^ However, burden of post-exposure quarantine for children exposed to infected children persisted amid rising incidence of infections.^7^ Test-to-stay approaches supplanted the conventional practice of post-exposure quarantine by frequent serial testing for 5-10 subsequent days and reduced the burden of post-exposure quarantine.^8–16^ Safety of test-to-stay approaches, expressed as equivalence in infections as compared to quarantine approaches, was addressed by several studies. ^8–16^ These reports are mostly restricted to screenings in schools and included the use of facemasks. Thus, evidence on safety of test-to-stay concepts in daycare facilities, without the use of facemasks, and in an age-group less capable of sticking to hygiene rules, is limited.

We previously reported on a city-wide SARS-CoV-2 Pool-polymerase chain reaction (PCR) screening in more than 700 daycare facilities in Cologne, Germany.^17^ After the reported period that included post-exposure quarantine measures, the screening was adapted to a test-to-stay approach and was continued for 30 weeks (September 2021-April 2022). Our study aimed to investigate the safety of this test-to-stay approach. We hypothesized that safety can be assumed if no substantial increase in frequencies of infections among children following the adaptation of the screening concept is detectable.

## Methods

### Ethics approval

All analyses were performed under a protocol approved by the institutional review board of the Medical Faculty of the University of Cologne (21-1358).

### Screening setting

The screening was implemented under the direction of the Youth Welfare Office of Cologne and accompanied by a team of members of the health authorities of Cologne, the University Hospital of Cologne, and a private diagnostic laboratory (“Labor Quade”). Participation in the screening program was voluntary. Sample collection was performed using the Lolli-Method as described previously (**Figure 1**).^17–19^ The Lolli-Method consists of sucking a nasopharyngeal swab for 30 seconds and subsequent testing in SARS-CoV-2 Pool-PCR. For generation of a pool of Lolli-swabs, each child of a daycare group placed a self-sampled Lolli-swab in a common collection tube. One tube containing all swabs of one respective group was tested in one PCR-reaction. When a Pool-PCR tested negative, all children of that pool were assumed to be SARS-CoV-2 negative. When the Pool-PCR tested positive, the respective children were re-tested individually on the next day in Single-PCRs. The identification of an infected child was followed by the isolation of this child. Children exposed to this child were quarantined up to 14 days during the quarantine approach. During the test-to-stay approach, exposed children were not quarantined but tested in Lolli-Single-PCRs for 5 subsequent days. If tested negative, they could continue to go to daycare.

**Figure 1:**
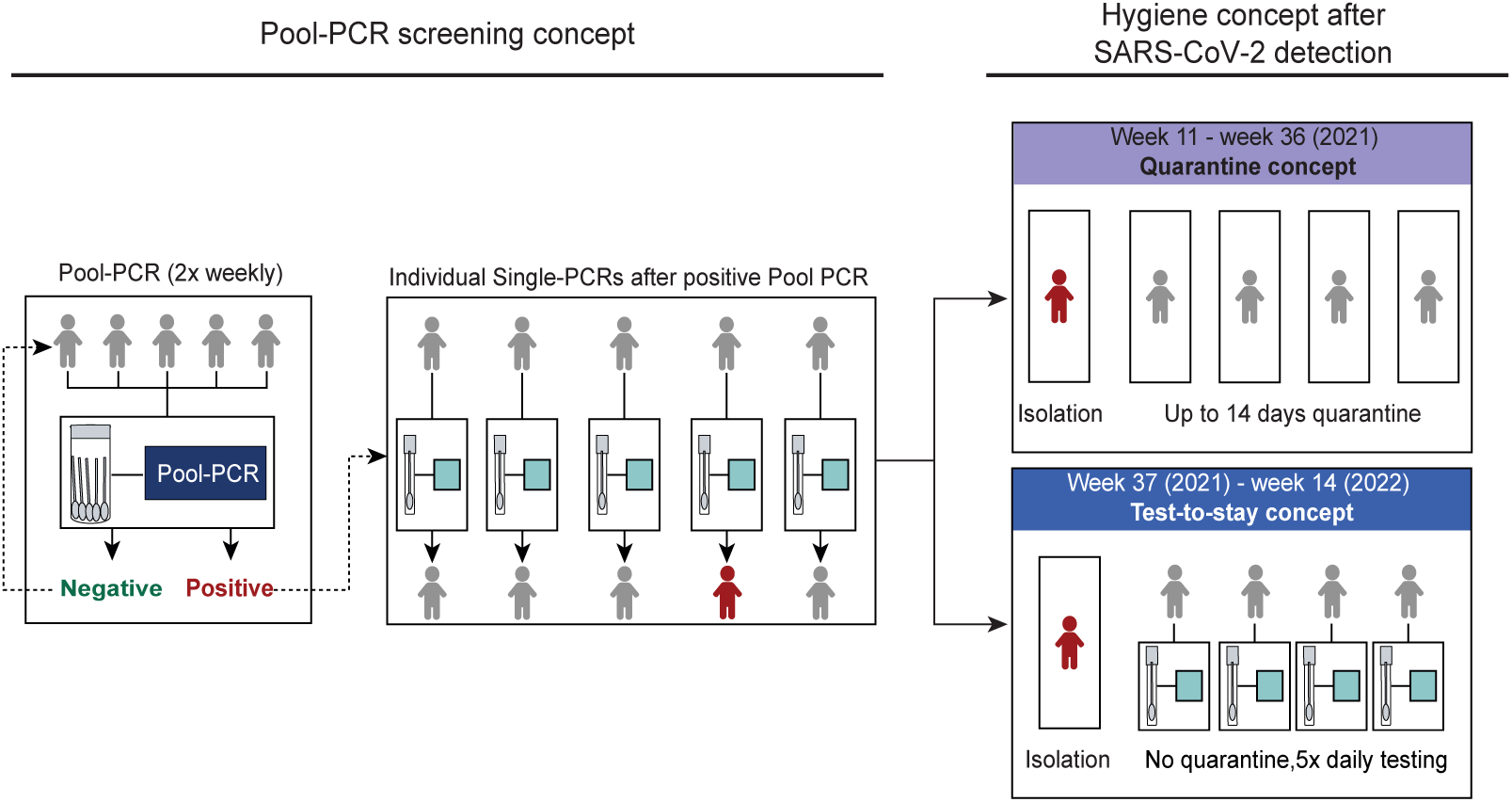
Overview on the screening concept and its specifications. Flowchart depicting the screening concept and both „Quarantine“ and „Test-to-stay“ approaches.

### Data sources

#### PCR test data

PCRs were performed at the Institute of Virology, University Hospital Cologne, and at Labor Quade. Labor Quade reported PCR test data to the Institute of Virology, containing the date of sampling, specimen (Pool- or Single-PCR), test result, daycare ID, and zip code of the daycare facility (**Supplemental Fig. 1**). During the roll-out of the screening (weeks 15-17 in 2021), daycare facilities were enrolled gradually, and the data collection system was under construction and misclassified the specimen of positive PCRs. These weeks were excluded from all analyses. Pool sizes were reported as weekly average pool size per daycare facility.

#### SARS-CoV-2 index cases and contact persons

The health authorities of Cologne provided data on newly reported SARS-CoV-2 infections (“index cases”) and contact persons recorded with a software developed for the documentation and case management of SARS-CoV-2 in Cologne.^20^ The data contained the date of the first SARS-CoV-2 detection or the date of the beginning and the end of the quarantine, age, sex, and the zip code of the place of residency (**Supplemental Fig. 1**).

#### Frequencies of SARS-CoV-2 variants of concern (VOCs) and 7-day incidence

Data on Germany-wide proportions of all reported sequences of VOCs were collected as part of a national molecular surveillance system and were published by the Robert Koch Institute (RKI).^21^ The 7-day incidence was extracted from RKI SurvStat@RKI2.0, an online tool for querying notifiable infectious diseases in Germany.^22^

#### Inhabitants and socioeconomic factors of Cologne

Aggregated data on the number of inhabitants in 2020 and socioeconomic factors were published online or provided by the Office for Urban Development and Statistics of Cologne.^23^

### Descriptive analysis

We reported counts and fractions of total and positive Pool-and Single-PCRs. Pool sizes were reported as median of the weekly mean pool sizes per daycare facility. Cumulative numbers of PCR analyses were estimated by multiplication of the counts of Pool-PCRs with the weekly median pool size and addition of the Single-PCRs. Distribution of positive Pool-PCRs among daycare facilities was reported as median positive Pool-PCRs per daycare facility. We calculated Spearman correlation coefficient to quantify the association between the weekly fraction of positive Pool-PCRs and the 7-day incidence.

We estimated the amount of quarantine that was avoided by the use of the test-to-stay approach with the following equations:

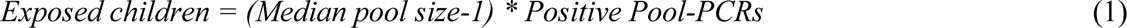

where *1* was subtracted from *Median pool* size to account for one assumed index case within one positive pool. *Positive Pool-PCRs* represents the number of positive Pool-PCRs during the test-to-stay approach. We assumed that all exposed children would have been quarantined in the absence of the test-to-stay approach.

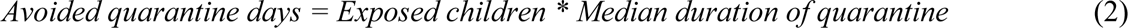

where *Median duration of quarantine* (days) referred to the screening approach involving quarantine after exposure.

To account for the variability of the individual parameters, we derived a variability range for the days of avoided quarantine by multiplying the first/third quartile of the pool size with the number of positive Pool-PCRs (equation 1) and the exposed children with the first/third quartile of the duration of quarantine (equation 2). The days of avoided quarantine per eligible child were estimated dividing the number of avoided quarantine days by the number of inhabitants in Cologne aged 2 to 6 years (n = 51,830), assuming that those were eligible for the screening.

### Regression discontinuity in time analysis

#### Outcome

The outcome for the RDiT was defined as the weekly count of index cases aged 2 to 6 years in Cologne divided by the weekly count of positive Pool-PCRs (**Supplemental Fig. 2**). We assumed that an infected child in a daycare facility would be detected in the screening and the corresponding positive Pool-PCR would contribute to the denominator of the outcome. We expected that secondary infections would occur in the daycare facility. The infected child and the secondary infections would be reported to the health authorities and contribute to the count of reported index cases aged 2 to 6 years in Cologne, which defined the numerator of the outcome. We hypothesized that the transition from the quarantine approach to the test-to-stay approach might affect the number of secondary infections and thus affect the outcome.

Analyses were restricted to weeks which did not meet the criteria of low-testing periods as we expected that in those, the denominator of the outcome would be underestimated (e.g., during roll-out of the screening or holiday seasons). We defined a low-testing period as a week in which less than 70% of the average weekly count of Pool-PCRs during the entire screening were performed.

#### RDiT framework

We followed current best practice for RDiT.^24–26^ The interruption was defined as the change of the test concept from a quarantine approach to a test-to-stay approach (week 37 in 2021). A lag-period of two weeks, in which observations were censored, followed the interruption, to account for the SARS-CoV-2 incubation period and to allow the newly introduced test concept to be fully implemented.^27^ We applied triangular kernel weights to assign higher weights to observations lying closer to the interruption. We modelled the outcome based on a gamma regression with the following equation:

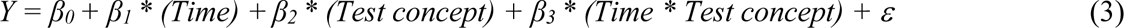

where *Y* is the outcome. *β_0_* is the intercept. *Time* is the running variable and given as weekly time units. *β_1_* reflects the slope before the interruption. *Test concept* is a dummy variable - 0 for the quarantine approach and 1 for the test-to-stay approach. The *β_2_*-coefficient describes the effect of the interruption on the outcome. The interaction term *Time * Test concept* allows for the slope of the regression line to differ on either side of the interruption (*β_3_)*. *ε* is the error term. Coefficients were exponentiated and interpreted on a multiplicative scale. Presence of autocorrelation was assessed with plots of functions of autocorrelation (acf) and partial autocorrelation (pacf).

#### Data-driven optimal bandwidth calculation

The width of the time period (“bandwidth”) drawn around the interruption is an important choice in RDiT.^24,25^ We determined a mean squared error (MSE)-optimal bandwidth, which minimizes the MSE of the regression fit.^28,29^ For the gamma regression model, we examined variations of the β_2_-coefficient across different bandwidth choices (1, 1.25, 1.5, 1.75, and 2 times the MSE-optimal bandwidth).

#### Robustness check

To assess the robustness of the gamma regression model, we extended the model specification given in equation (3) to a second-order polynomial regression instead of a linear regression (**Supplemental Information, Appendix A**).

#### Testing continuity of baseline covariates

RDiT requires continuity of baseline covariates across the interruption.^24,25^ This verifies that changes in the outcome are attributable to the defined interruption and not to coincident changes of covariates. Continuity was assessed with help of the RDiT framework by applying local linear regressions. Assuming normal distribution, we modelled the median pool size, the fraction of the Delta variant and the 7-day incidence.

#### Sensitivity analyses

We included the 7-day incidence, the seasons, and the fraction of the respective SARS-CoV-2 variant in Germany in the gamma regression model (**Supplemental Information, Appendix A**). We ran stratified analyses for districts with low and middle/high socioeconomic status (SES). For stratification, we used a previously described index of the SES of the neighborhoods of Cologne (**Supplemental Information, Appendix B**).^30^

### Software

Statistical analyses were performed using Microsoft Excel for Mac (v.14.7.3.), Prism 9.0 (GraphPad) and R within RStudio (v. 2023.03.0+386) and additional R packages.^31–33^

## Results

### Implementation of the screening program

The screening was conducted from March 2021-April 2022 in Cologne, Germany (1.1 million inhabitants). All daycare facilities in Cologne and 51,830 children aged 2 to 6 years were eligible to participate twice weekly. The quarantine approach was conducted from week 11 to 36 (2021) and was substituted by the test-to-stay approach from week 37 (2021) to week 14 (2022) (**Table 1**, **Figure 2**). 714 daycare facilities participated in the screening. During the screening, four VOCs emerged (Alpha, Delta, BA.1, and BA.2) and the SARS-CoV-2 7-day incidence in Cologne ranged from 8.67 infections per 100,000 inhabitants in week 25 (2021) to 2,573 in week 9 (2022) (**Supplemental Fig. 3**).

**Figure 2:**
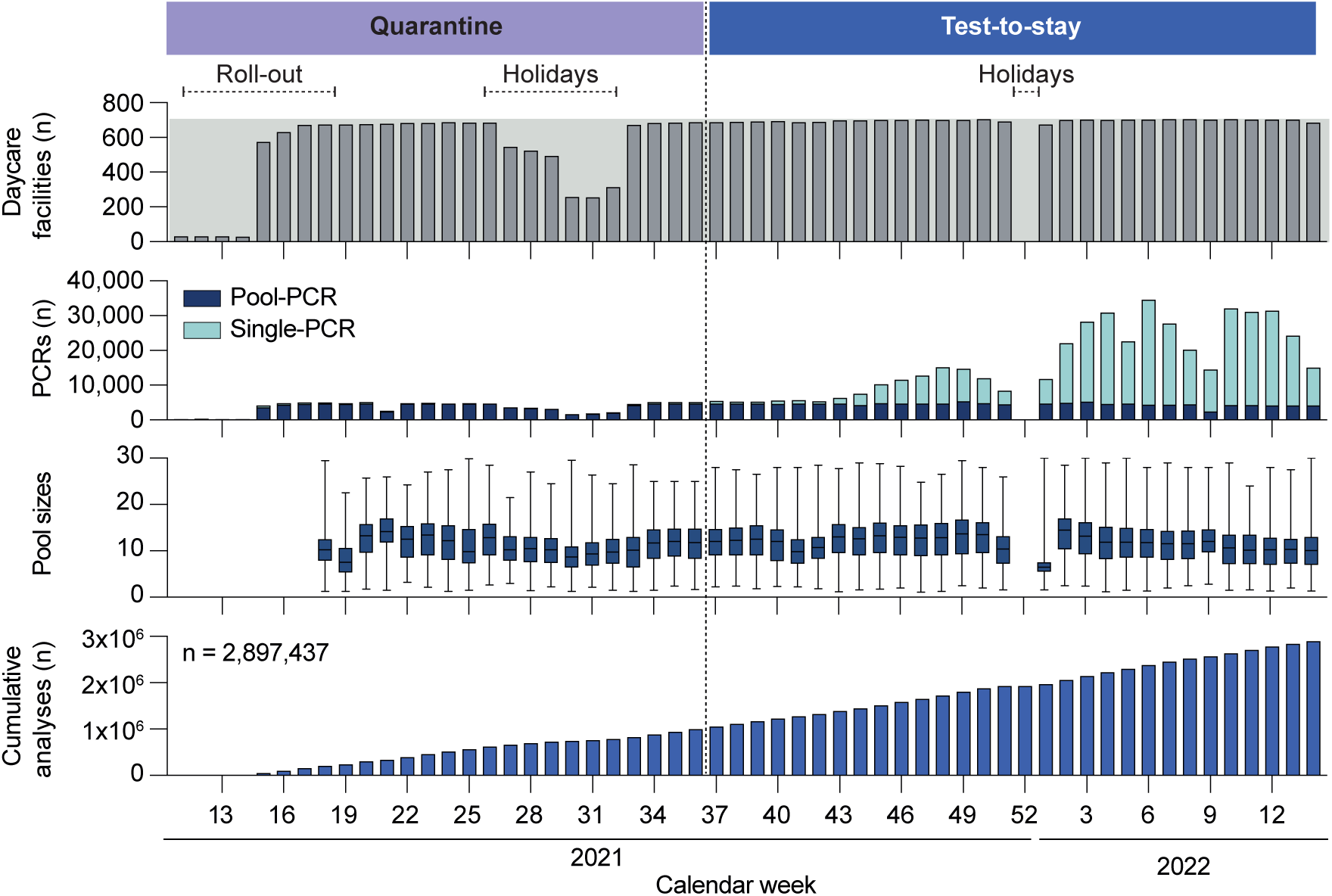
Implementation of the screening concept in daycare facilities. The number of tested daycare facilities, performed PCRs, median pool sizes and cumulative number of performed analyses are stratified by calendar week. The horizontal lines in the Box-Whisker-Plot indicate the medians, the lines at the top and at the bottom of the boxes indicate first and third quartiles and the error bars represent minimum and maximum pool sizes. Data on pool sizes were not available during the roll-out.

**Table 1:** Summary of the screening program.

| **Characteristics** | **Quarantine approach\***<br>n (%) | **Test-to-stay approach**<br>n (%) | **Entire screening**<br>n (%) |
| --- | --- | --- | --- |
| **Duration** |  |  |  |
| **Time period** | Week 11 2021 - week 36 2021 | Week 37 2021 - week 14 2022 | Week 11 2021 - week 14 2022 |
| **Weeks** | 26 | 30 | 56 |
| **Daycare facilities** |  |  |  |
| **Total** | 694 | 712 | 714 |
| **Pool-PCRs** |  |  |  |
| **Total** | 87,856 | 132,029 | 219,885 |
| **Positive\*** | 142 (0.16) | 6298 (4.77) | 6440 (2.93) |
| **Single-PCRs** |  |  |  |
| **Total** | 5,704 | 346,601 | 352,305 |
| **Positive\*** | 111 | 12,343 | 12,454 |
| **Index cases (2-6 years in Cologne)** |  |  |  |
| **Total** | 1,203 | 16,005 | 17,208 |

\*During the roll-out of the screening (weeks 15-17 in 2021), daycare facilities were enrolled gradually, and the data collection system was under construction and misclassified the specimen of positive PCRs. Thus, these weeks were excluded from all analyses.

219,885 Pool-PCRs and 352,305 Single-PCRs were performed. Median of weekly mean pool sizes per daycare facility was 11.5 (IQR: 8-14.6) swabs per pool (**Supplemental Fig. 4**). Approximately 2,897,437 SARS-CoV-2 analyses were performed in total (**Table 1**, **Figure 2**). 6,440 (2.93%) of the Pool-PCRs tested positive (**Table 1)**. The median number of positive Pool-PCRs per daycare facility was 9 (IQR: 5-12) (**Supplemental Fig. 5A**). The weekly fraction of positive Pool-PCRs ranged from 0.0% in weeks 24-27 (2021) to 12.44% in week 9 (2022) (**Figure 3A, Supplemental Fig. 5B**) and correlated strongly with the total SARS-CoV-2 7-day incidence in Cologne (r_s_ = 0.96, CI: 0.94-0.98; **Figure 3B**). 12,454 of the Single-PCRs tested positive. During the entire screening, the health authorities in Cologne reported 17,208 index cases among children aged 2 to 6 years **(Table 1**, **Figure 3A**). Subsequently, our estimation suggests that the screening identified 72.4% of all reported index cases aged 2 to 6 years in Cologne.

**Figure 3:**
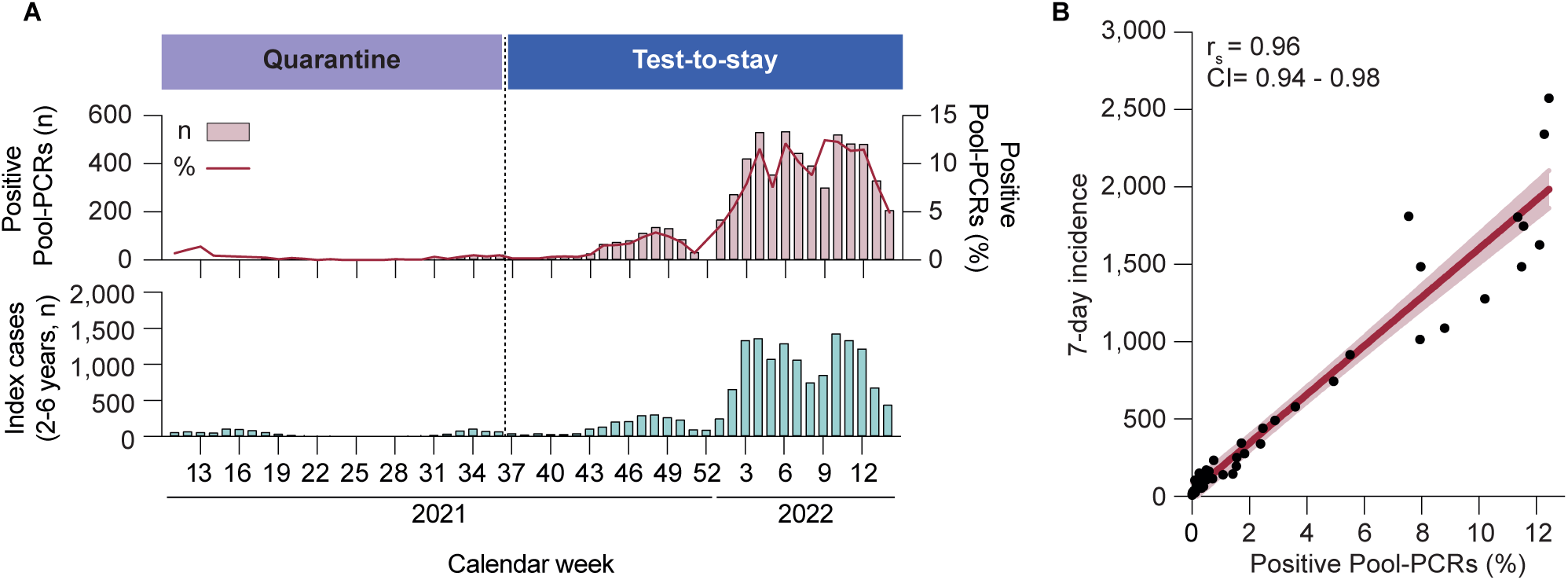
Detection of SARS-CoV-2 infections in daycare facilities. **A**, the number and fraction of positive Pool-PCRs and the number of reported index cases (2-6 years) are stratified by calendar week. During the roll-out of the screening (weeks 15, 16, 17 in 2021) the data collection system misclassified the specimen (Pool or Single-PCR) of positive PCRs. Thus, data on positive PCRs are not available for these weeks. **B**, Spearman correlation between 7-day incidence in Cologne and fraction of positive Pool-PCRs. Each black dot represents one week. 95% CI is indicated by the bright red area.

During the test-to-stay approach, 6,298 Pool-PCRs tested positive (**Table 1**). The median duration of quarantine after exposure to SARS-CoV-2 of children aged 2 to 6 years in Cologne was 10 days (IQR: 8;12) (**Supplemental Fig. 6**). Consequently, we estimated that 661,290 days (variability range: 352,688-1,027,833) spent in quarantine were avoided during the test-to-stay approach. This translated to 13 days (variability range: 7-20) of avoided quarantine per eligible daycare child during a period of 30 weeks (**Figure 4**).

**Figure 4:**
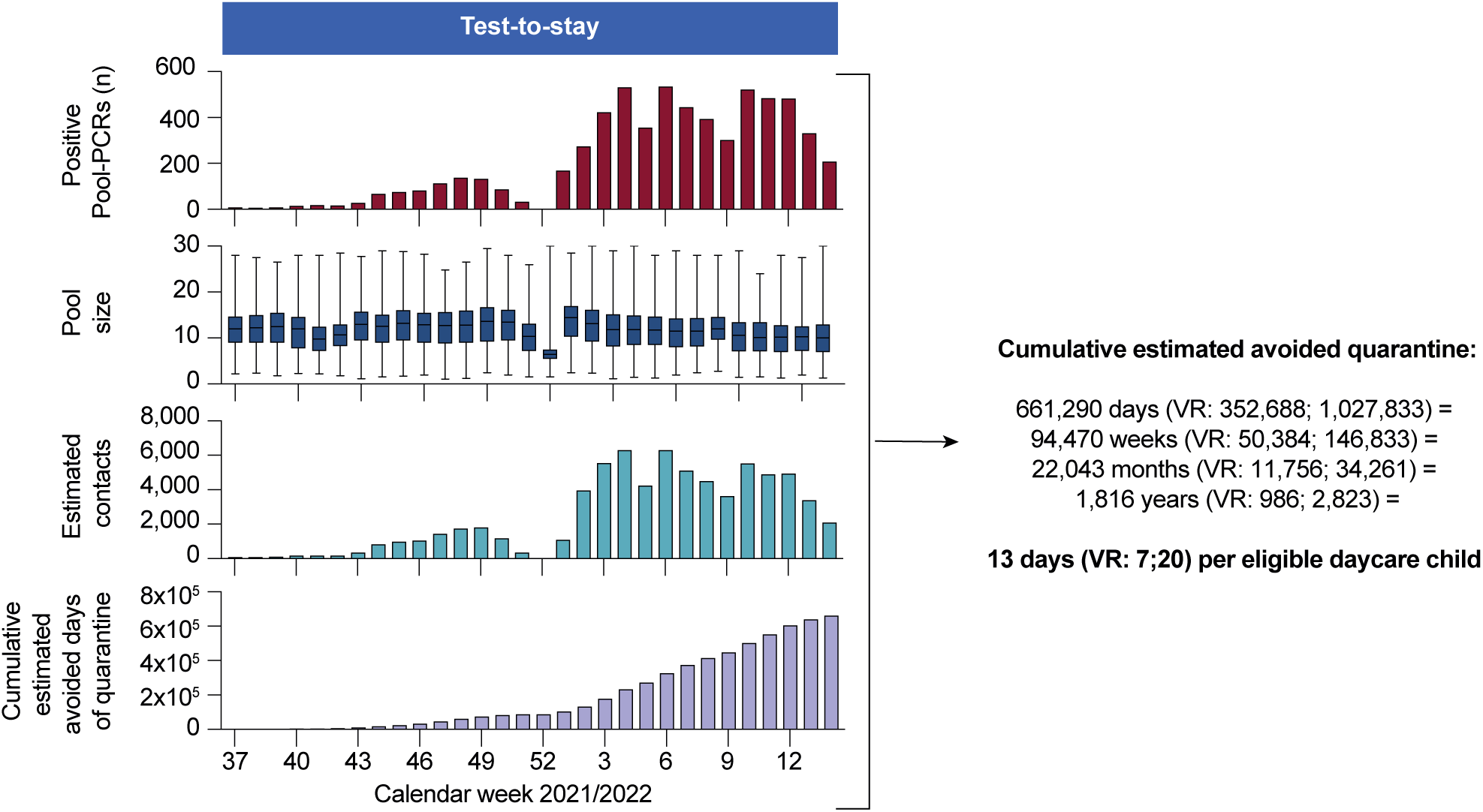
Estimation of quarantine avoidance. The number of positive Pool-PCRs, pool sizes, estimated contact persons and cumulative estimated avoided days of quarantine are stratified by calendar week. The horizontal lines in the Box-Whisker-Plot indicate the medians, the lines at the top and at the bottom of the boxes indicate first and third quartiles and the error bars represent minimum and maximum pool sizes.

### Safety of the test-to-stay approach

Applying an RDiT analyses, we aimed to assess the safety of the test-to-stay approach in comparison to the quarantine approach with the underlying causal framework depicted in a directed acyclic graph (**Supplemental Fig. 7, Supplemental Table 1**).^25^ We first incorporated all observations in the gamma regression model (”<u>global</u> gamma regression model”) which yielded an exponentiated β_2_-coefficient of 0.74 (95% CI: 0.52;1.06) (**Figure 5A**). This translated to 26% less index cases per positive Pool-PCR attributable to the change of the test concept and indicated no substantial increase in infections among children. Visualization of the acf and pacf showed no clear presence of autocorrelated residuals (**Supplemental Fig. 8**). To address biases arising from potential unobserved confounders that are not in proximity of the interruption, we gradually narrowed the underlying bandwidth of the global gamma regression model in a sequence of local gamma regressions (**Figure 5B, Supplemental Fig. 9**). Overall, the main result of the global gamma regression model was not sensitive to the choice of the bandwidth. The observed decrease in precision for shorter bandwidths is a standard finding in RDiT.^24,25^ As robustness check, we verified that the result of the global gamma regression model was similar when using a second-order polynomial specification instead of a linear regression (**Supplemental Fig. 10**).

**Figure 5:**
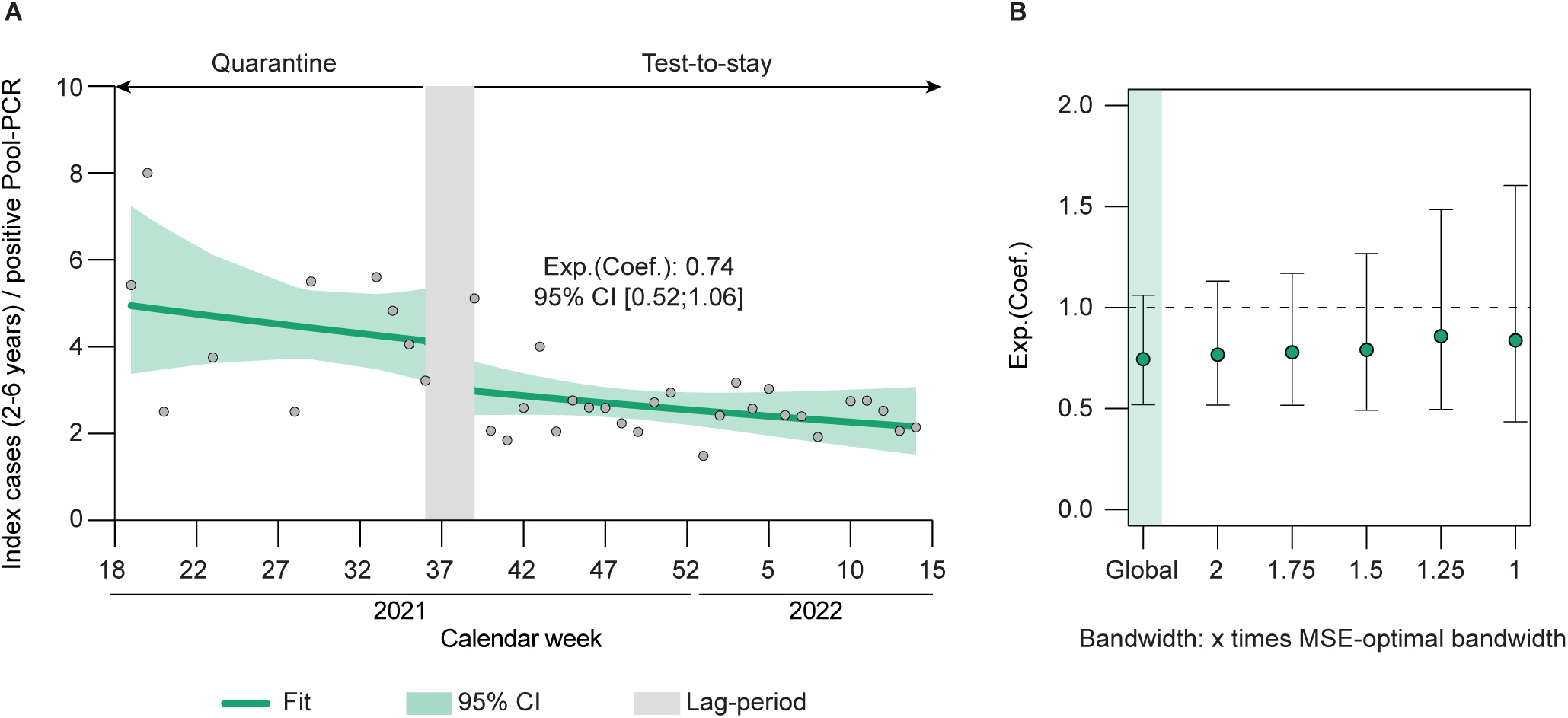
Assessment of safety of the test-to-stay approach: Regression discontinuity in time analysis (RDiT) **A**, RDiT global gamma regression model. Model fit, 95% CI and the lag period are indicated in their respective color. **B**, Exponentiated coefficient estimates of local gamma regressions with distinct bandwidths. Error bars indicate 95% CI.

### Testing continuity of baseline covariates

The weekly median pool size (β_2_ = -0.89, CI: -4.12;3.23) and the share of the Delta variant (β_2_ = -0.57, CI: -1.42;0.28) showed continuity among the interruption. The 7-day incidence did not meet the continuity assumption (β_2_ = -147, CI: -244;-49) (**Supplemental Fig. 11**).

### Sensitivity analyses

As 7-day incidence showed discontinuity across the interruption, we included it as predictor in the global gamma regression model. To account for a potential impact of seasonality, we included the seasons in the global gamma regression model. As we could not rule out a role as confounder or competing exposure of the SARS-CoV-2 variant, we included it as predictor in the global gamma regression model. Accounting for these covariates in separate global models, we detected no substantial differences of the corresponding effect estimates or their respective precision (**Supplemental Fig. 12**). Stratification by SES indicated differences between the β_2-_ coefficients of both regressions (high/middle: exp. β_2_ = 0.57, CI: 0.37;0.85 and low: exp. β_2_ = 1.29, CI: 0.73;2.24). However, in both strata, there was no indication of an increase in infection frequency (**Supplemental Fig. 13**).

## Discussion

This study provided evidence that suggest safety of a SARS-CoV-2 Pool-PCR test-to-stay screening in daycare facilities as compared to a post-exposure quarantine concept. We analyzed one of the most comprehensive test-to-stay screenings in daycare facilities worldwide.^34–37^ This screening was one of the earliest attempts to disestablish the use of quarantine of daycare children in Germany. Our estimation of the impact of the test-to-stay concept on quarantine avoidance showed a substantial reduction in the time spent in quarantine. Our RDiT analyses did not indicate evidence of a discernible increase in infections after quarantine measures were discontinued.

Our main results on safety are in line with previous reports on test-to-stay approaches.^8–16^ However, those studies were primarily conducted in schools with children and adolescents older than 6 years, and they incorporated the use of facemasks. In contrast, the here analyzed screening was conducted in the absence of the use of facemasks and in an age-group less capable of sticking to hygiene rules. Furthermore, the mentioned studies were conducted during the predominance of one respective SARS-CoV-2 VOCs.^8–16^ In contrast, our analyses spanned a period during which four distinct SARS-CoV-2 VOCs have emerged (Alpha, Delta, BA.1, and BA.2). These variants show distinct virological features that could affect the effectiveness of infection control measures (e.g., incubation period or basic reproduction number).^38–41^ We show that safety of the test-to-stay screening was not affected by these VOCs. Finally, previous studies mainly report on short-term observations with rather small variations in the incidence, whereas our analyses encompass 13 months with low-and high incidence periods, providing more generalizable evidence.

Lolli-swabs collect mainly saliva which has been shown to be a valid specimen for the detection of other respiratory viruses.^42–47^ Thus, a test-to-stay approach involving self-sampling of saliva in daycare facilities might be considered an alternative to quarantine in a future epidemic or pandemic scenario of other respiratory viruses. It might contribute to an age-appropriate somatic and physical development.

One limitation of this study is that the ethical and political circumstances during the COVID-19 pandemic did not justify addressing the research question by a randomized controlled trial, emerging from the initial screening with a quarantine approach. Thus, the quasi-experimental study design differs from the ideal experimental design. Second, as we analyzed aggregated data, we cannot deduce any causal statements on the level of individual children. Further individualization of the data was not possible due to the challenge of matching index cases with their corresponding positive Pool-PCRs. Third, the risk of bias due to unmeasured confounders needs to be acknowledged. It might comprise changes in data collection practices of the health authorities or in test indication for SARS-CoV-2 testing beyond the screening. However, we assume that most index cases were constantly reported and recorded, as the screening detected most of all notified index cases among children. Furthermore, we assumed that the risk of confounding increases with increasing distance from the interruption and applied kernel weights and different bandwidths to address this risk.

## Conclusion

The test-to-stay approach substantially reduced the use of quarantine after SARS-CoV-exposure. There was no indication of a relevant increase in infections among children with this approach. This highlights it as a safe alternative to quarantine after exposure to SARS-CoV-2 in daycare. Test-to-stay approaches could prove valuable as infection control measures after exposure to emerging respiratory pathogens in daycare during future outbreaks.

## Data Availability

Deidentified PCR test data (including data dictionaries) will be made available, in addition to study protocols, the statistical analysis plan, and the code for analysis. The data and code will be made available upon publication to researchers who provide a methodologically sound proposal for use in achieving the goals of the approved proposal. Proposals should be submitted to.

## Acknowledgements

The authors thank all children and staff in tested daycare facilities for support and participation. We thank Moritz Lorenz and Paula Lorenz for supporting the development of the Lolli-Method. We thank Sascha Nickel and Anne Fries as well as all staff members of their daycare facilities for their impetus for the development of the Lolli-Method and screening adaptions. We thank all members of the Institute of Virology, University Hospital Cologne. We thank all staff of Labor Quade for conducting the screening and providing data. We thank Carsten Tschirner (IExcelU) for supporting data visualization and Stephan Glaremin (Amt für Jugend, Arbeit und Soziales der Stadt Düsseldorf) for administrative support during implementation of the screening in Cologne. We thank Janna Seifried and Sindy Böttcher (Robert Koch Institute) for continuous support and discussions. We thank Stefan Konigorski and Mayram Ganji (Berlin School of Public Health) for statistical support and discussion.

## Abbreviations

COVID-19: Coronavirus disease 2019
SARS-CoV-2: severe acute respiratory syndrome coronavirus type 2
PCR: polymerase chain reaction
RDiT: regression discontinuity in time
MSE: mean squared error
SES: socioeconomic status
IQR: inter-quartile range
RKI: Robert Koch Institute

