## Supplement for "Safety of SARS-CoV-2 test-to-stay in daycare: a regression discontinuity in time analysis"

**Supplemental Information**

### Data sources

**Supplemental Figure 1**

Overview on analyzed data on PCRs of the screening and index cases in Cologne

### Methods

**Supplemental Figure 2**

Conceptual framework of the main outcome of the RDiT analyses

**Appendix A**

Regression discontinuity in time

**Appendix B**

Socioeconomic status of city districts

### Results

**Supplemental Figure 3**

SARS-CoV-2 VOCs and 7-day incidence during the screening

**Supplemental Figure 4**

Overview on analyzed data on pool sizes of the screening

**Supplemental Figure 5**

Further description of detected SARS-CoV-2 infections in daycare facilities

**Supplemental Figure 6**

Overview on analyzed data on duration of quarantine

**Supplemental Figure 7**

Directed acyclic graph depicting the causal framework of the RDiT

**Supplemental Table S1**

Assumptions in the causal paths of the directed acyclic graph with their rationale and references, if available

**Supplemental Figure 8**

Assessment of autocorrelation of the residuals of the RDiT

**Supplemental Figure 9**

RDiT models with different bandwidth specifications

**Supplemental Figure 10**

RDiT analysis using global squared regression instead of global linear regression

**Supplemental Figure 11**

Testing continuity of baseline covariates

**Supplemental Figure 12**

Sensitivity analyses 1

**Supplemental Figure 13**

Sensitivity analyses 2

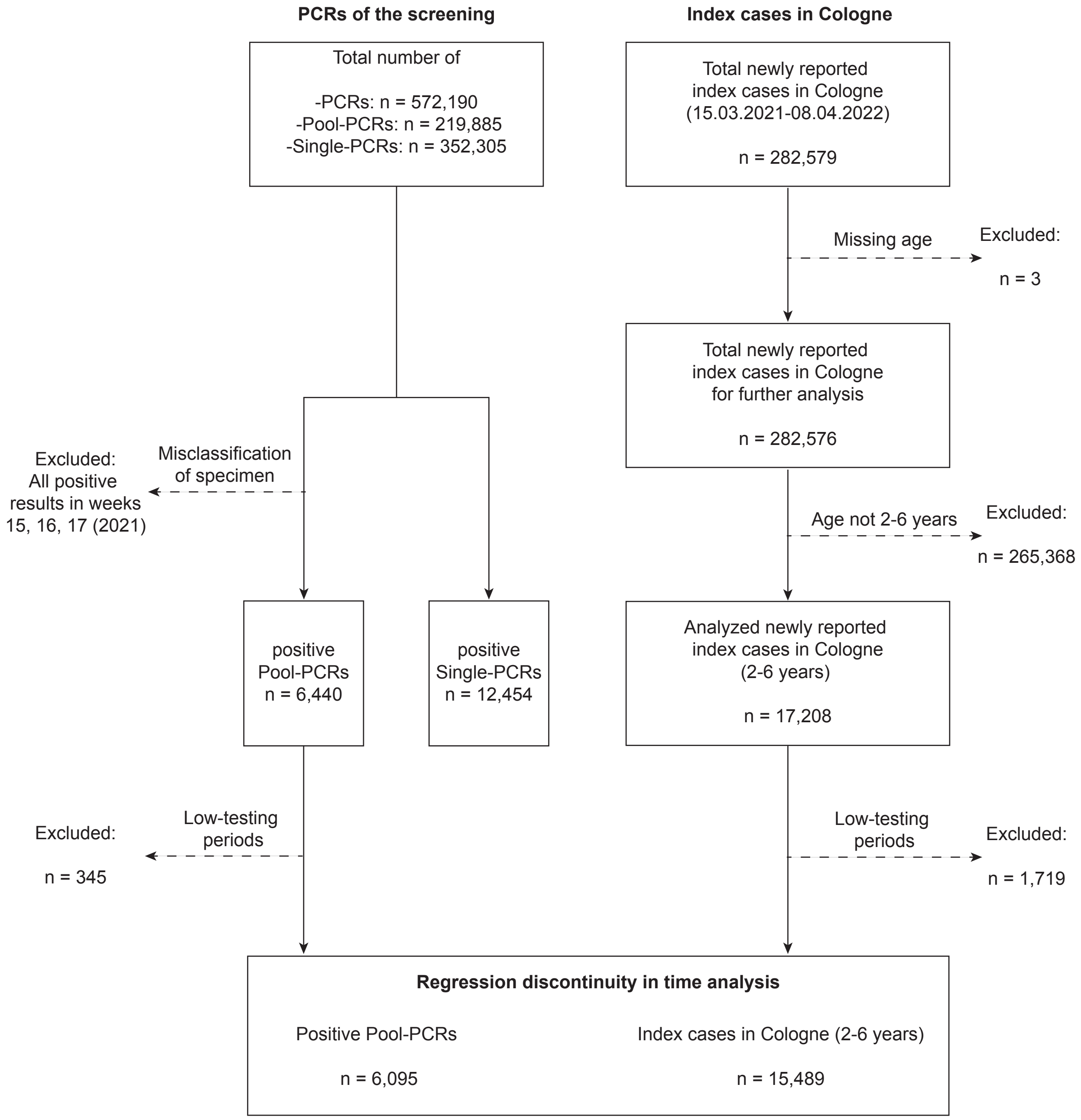


**Supplemental Figure 1: Overview on analyzed data on PCRs of the screening and index cases in Cologne**

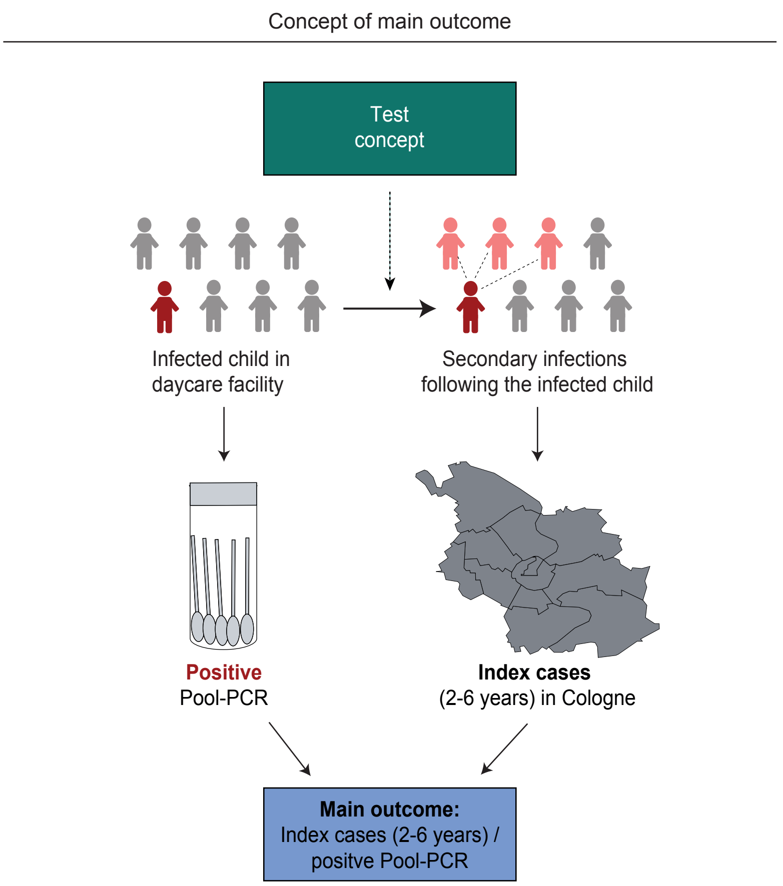


**Supplemental Figure 2: Conceptual framework of the main outcome of the RDiT analyses**

**Supplementary Methods, Appendix A:** Regression discontinuity in time (RDiT)

### Robustness check

To assess the robustness of the global gamma regression model, we extended the model specification given in equation (3) to a second-order polynomial regression instead of a linear regression, using the following equation:

$$Y = \beta_0 + \beta_1 * (Time) + \beta_2 * (Test\ concept) + \beta_3 * (Time * Test\ concept) + \beta_4 * (Time^2) + \beta_5 * (Time^2 * Test\ concept) + \varepsilon \quad (4)$$

### Sensitivity analyses

To account for the 7-day incidence, the season, and the SARS-CoV-2 variant and the respective influence on the effect estimate, we modified the model predicting the outcome specified by equation (3) as follows:

$$Y = \beta_0 + \beta_1 * (Time) + \beta_2 * (Test\ concept) + \beta_3 * (Time * Test\ concept) + \beta_4 * (7\text{-}day\ incidence), \quad (5)$$

where *7-day incidence* was used as continuous variable, and

$$Y = \beta_0 + \beta_1 * (Time) + \beta_2 * (Test\ concept) + \beta_3 * (Time * Test\ concept) + \beta_4 * (Spring) + \beta_5 * (Summer) + \beta_6 * (Autumn) + \beta_7 * (Winter), \quad (6)$$

where *Spring*, *Summer*, *Autumn*, and *Winter* were used as indicator variables reflecting the respective season, and

$$Y = \beta_0 + \beta_1 * (Time) + \beta_2 * (Test\ concept) + \beta_3 * (Time * Test\ concept) + \beta_4 * (Alpha) + \beta_5 * (Delta) + \beta_6 * (BA.1) + \beta_7 * (BA.2), \quad (7)$$

where *Alpha*, *Delta*, *BA.1*, and *BA.2* are continuous variables, reflecting the fraction of the respective SARS-CoV-2 variant in Germany.

**Supplementary Methods, Appendix B:** Socioeconomic status of city districts

Neuhann et al. have previously assigned a socioeconomic index to the neighborhoods of Cologne.[1] They based their socioeconomic index on three factors: i) the proportion of the population of the districts with migration background, ii) the proportion of households in need of assistance (benefit units) among all private households, and iii) the proportion of inhabitants that are unemployed. Since this socioeconomic index was originally developed for the 86 neighborhoods, we adapted it to categorize the 9 districts of Cologne, which include multiple neighborhoods and divided the city districts by categories of SES (high, middle, and low). To this end, proportions of migration background, benefit units, and unemployment were normalized with 0 reflecting the minimum and 1 the maximum value of each characteristic. Subsequently, the normalized values of the respective characteristic of the districts were summed with equal weights (weight = 0.33) to compute a numeric index. Finally, this normalized index for SES was divided by three percentiles to assign high, middle, and low socioeconomic status to the first, second, and third percentile, respectively.

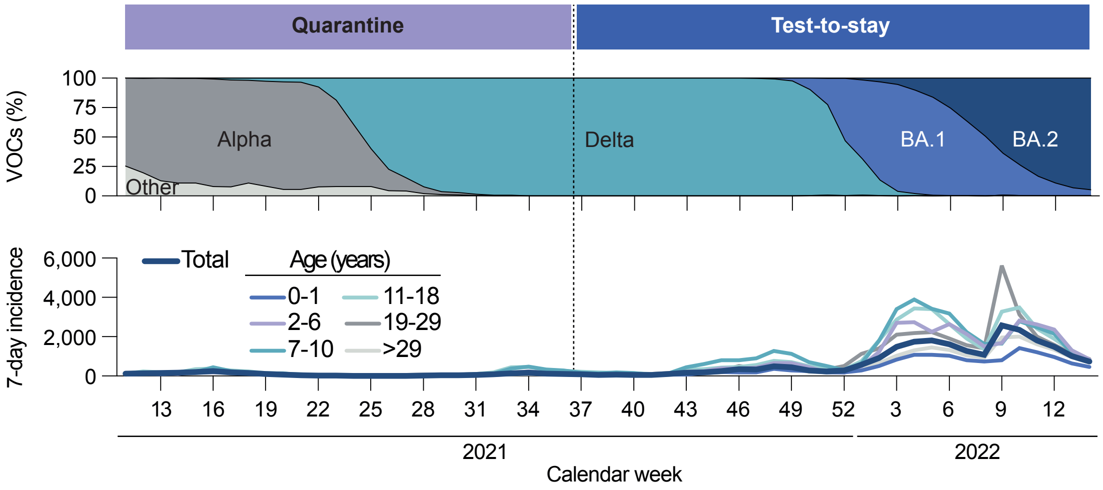


**Supplemental Figure 3: SARS-CoV-2 VOCs and 7-day incidence during the screening**

The frequencies of variants of concern and the 7-day incidence of different age groups, according to data published by the Robert Koch Institute, are stratified by calendar week.

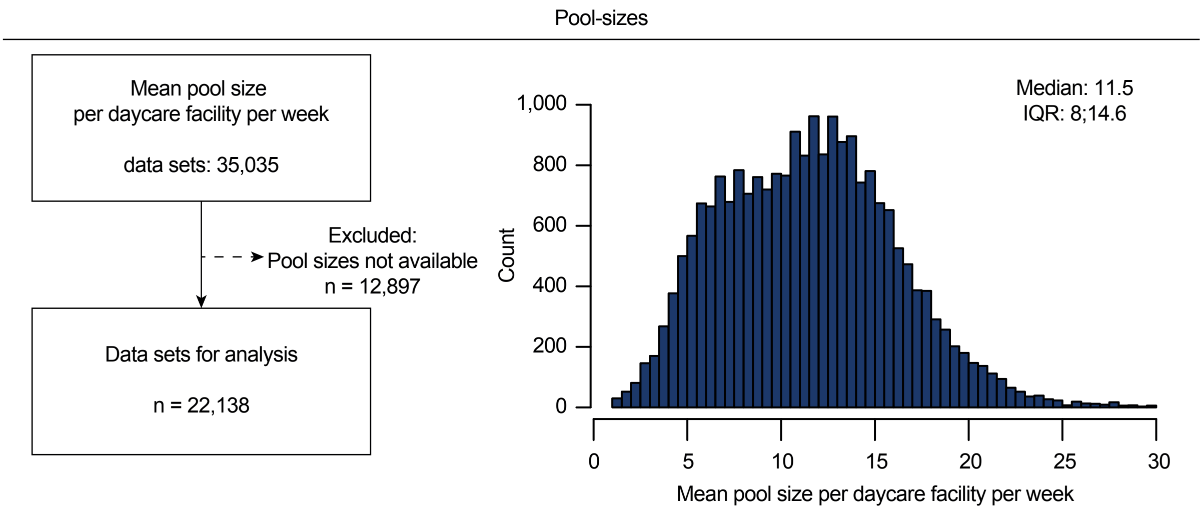


**Supplemental Figure 4: Overview on analyzed data on pool sizes**

The flowchart depicts the analyzed data on pool sizes (left). The histogram illustrates the distribution of the weekly mean pool sizes per daycare facility (right).

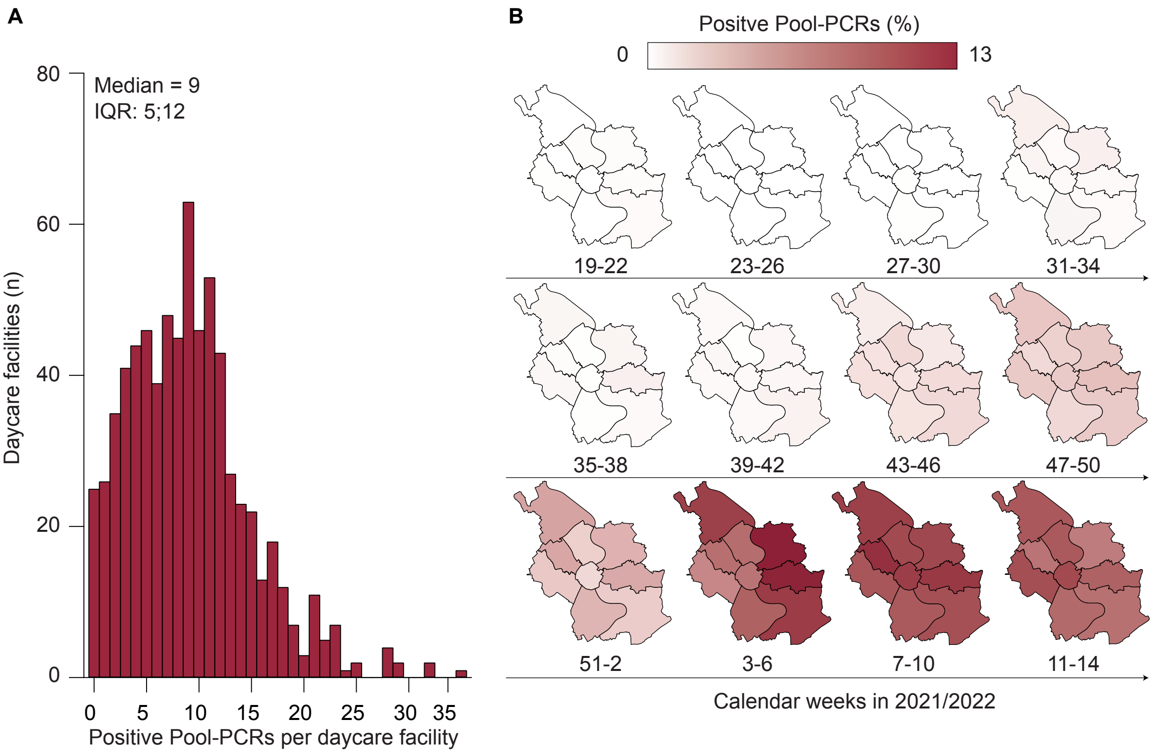


**Supplemental Figure 5: Further description of detected SARS-CoV-2 infections in daycare facilities**

**A**, the counts of daycare facilities are stratified by corresponding counts of positive Pool-PCRs per facility during the screening.
**B**, maps of Cologne depicting the fraction of positive Pool-PCRs per 4 weeks and city district.

Duration of quarantine

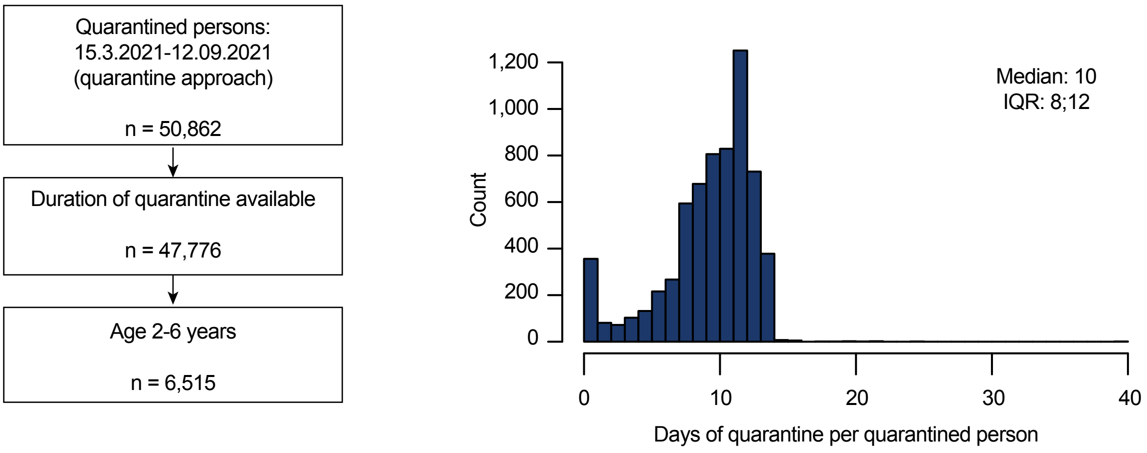


**Supplemental Figure 6: Overview on analyzed data on quarantine duration**

The flowchart depicts the analyzed data on duration of quarantine during the quarantine approach of the screening (left). The histogram illustrates the distribution of the duration of quarantine (right).

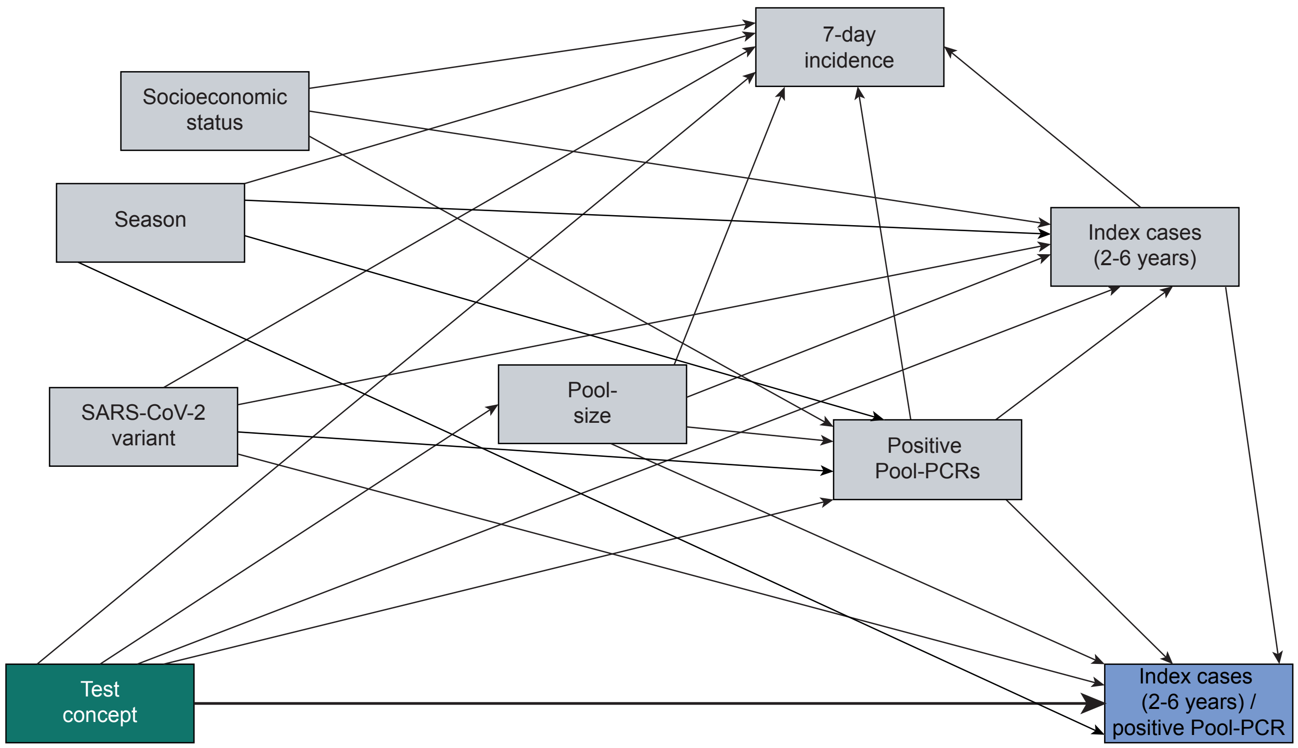


**Supplemental Figure 7: Directed acyclic graph depicting the causal framework of the RDiT**

### Supplemental Table 1: Assumptions in the causal paths of the directed acyclic graph (DAG) with their rationale and references, if available

| Cause | Direction | Effect | Rationale | Reference if available |
|---|---|---|---|---|
| Test concept | → | Index cases (2-6 years) / positive Pool-PCRs | If the change of the test concept is followed by a change in the number of secondary infections in daycare facilities, the ratio of index cases (2-6 years) / positive Pool-PCRs is altered. | NA |
| Test concept | → | Positive Pool-PCRs | Changes in the test concept may lead to changes in infection dynamics which could be mirrored by changes in the number of positive Pool-PCRs. | NA |
| Test concept | → | Index cases (2-6 years) | Changes in the test concept may lead to changes in infection dynamics which could be mirrored by changes in the number of reported index cases (2-6 years) in Cologne. | NA |
| Test concept | → | Pool size | Changes in the test concept may be followed by changes in pool sizes. | NA |
| Test concept | → | 7-day incidence | Changes in the test concept may lead to changes in infection dynamics which could be mirrored by changes in the 7-day incidence in Cologne. | NA |
| SARS-CoV-2 variant | → | Positive Pool-PCRs | Properties of newly emerged SARS-CoV-2 variants (e.g., basic reproduction number, incubation period, clinical presentation) may have an influence on infection dynamics in daycare facilities and by that change the number of positive Pool-PCRs. | 2 |

**Supplemental Table 1 cont.**

| Cause | Direction | Effect | Rationale | Reference if available |
|---|---|---|---|---|
| SARS-CoV-2 variant | → | Index cases (2-6 years) | Properties of newly emerged SARS-CoV-2 variants (e.g., basic reproduction number, incubation period, clinical presentation) may have an influence on infection dynamics in daycare facilities and by that may change the number of index cases (2-6 years) in Cologne. | 2,3,4,5 |
| SARS-CoV-2 variant | → | Index cases (2-6 years) / positive Pool-PCRs | Properties of newly emerged SARS-CoV-2 variants (e.g., basic reproduction value, incubation number, clinical presentation) may have an influence on infection dynamics in daycare facilities and by that may alter the ratio of index cases (2-6 years) / positive Pool-PCRs. | 2,3,4,5 |
| SARS-CoV-2 variant | → | 7-day incidence | Properties of newly emerged SARS-CoV-2 variants (e.g., basic reproduction number, incubation period, clinical presentation) may have an influence on infection dynamics in daycare facilities and by that may change the 7-day incidence in Cologne. | 3,4,5 |
| Season | → | 7-day incidence | The seasonality of SARS-CoV-2 might influence the 7-day incidence in Cologne. | 6,7 |
| Season | → | Index cases (2-6 years) | The seasonality of SARS-CoV-2 might influence the number of index cases (2-6 years) in Cologne. | 6,7 |

**Supplemental Table 1 cont.**

| Cause | Direction | Effect | Rationale | Reference if available |
|---|---|---|---|---|
| Season | → | Positive Pool-PCRs | The seasonality of SARS-CoV-2 might influence the number of positive Pool-PCRs. | 6,7 |
| Season | → | Index cases (2-6 years) / positive Pool-PCRs | The seasonality of SARS-CoV-2 might influence the main outcome. | 6,7 |
| Socioeconomic status (SES) | → | 7-day incidence | SARS-CoV-2 infections are associated with the socioeconomic status. This might be mirrored by differences in the 7-day incidence in districts with different SES. | 2,8,9 |
| Socioeconomic status (SES) | → | Index cases (2-6 years) | SARS-CoV-2 infections in children are associated with the socioeconomic status. This might be mirrored by differences in the number of indey cases (2-6 years) in districts with different SES. | 2,10 |
| Socioeconomic status (SES) | → | Positive Pool-PCRs | SARS-CoV-2 infections are associated with the socioeconomic status. This might be mirrored by differences in the number of positive Pool-PCRs in districts with different SES. | 2,11,12 |
| Pool size | → | Index cases (2-6 years) / positive Pool-PCRs | The pool size has an influence on the positivity of the Pool-PCRs as it might decrease with decreasing pool size. By that, changes in the pool sizes might influence the ratio of index cases (2-6 years) / positive Pool-PCRs. | NA |

**Supplemental Table 1 cont.**

| Cause | Direction | Effect | Rationale | Reference if available |
|---|---|---|---|---|
| Positive Pool-PCRs | → | 7-day incidence | It is assumed that the screening detects asymptomatic children that would have been tested later if the screening would not have taken place. Thus, the number of positive Pool-PCRs is one cause of the 7-day incidence in Cologne and not vice versa. | NA |
| Positive Pool-PCRs | → | Index cases (2-6 years) | It is assumed that the screening detects asymptomatic children that would have been tested later if the screening would not have taken place. Thus, the number of positive Pool-PCRs is one cause of the number of index cases (2-6 years) in Cologne and not vice versa. | NA |
| Positive Pool-PCRs | → | Index cases (2-6 years) / positive Pool-PCRs | The number of positive Pool-PCRs is defined as denominator of the main outcome. | NA |
| Index cases (2-6 years) | → | Index cases (2-6 years) / positive Pool-PCRs | The number of index cases (2-6 years) is defined as numerator of the main outcome. | NA |

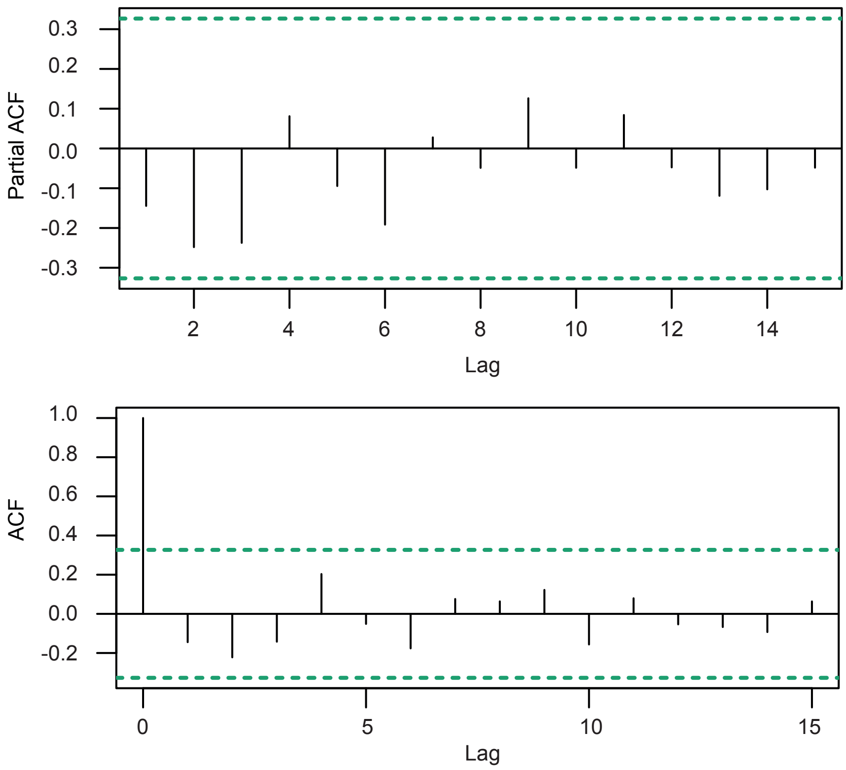


**Supplemental Figure 8: Assessment of autocorrelation of the residuals of the RDiT**

The partial autocorrelation function is plotted in the top panel. The autocorrelation function is plotted in the bottom panel. There is no indication of autocorrelation.

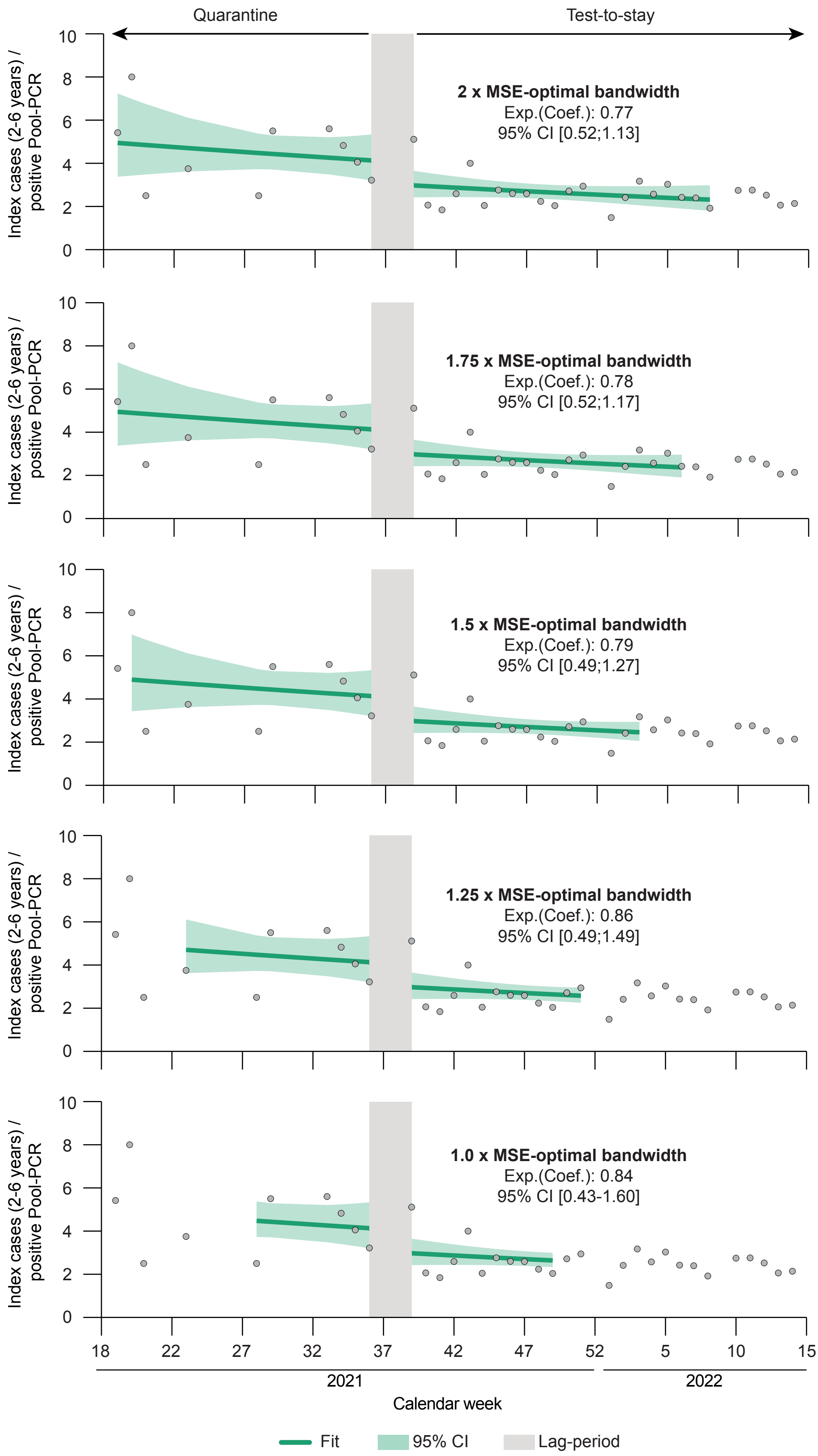


**Supplemental Figure 9: RDiT models with different bandwidth specifications**

The bandwidths are defined according to data-driven MSE-optimal bandwidth selection. The exponentiated coefficients are interpreted on a multiplicative scale.

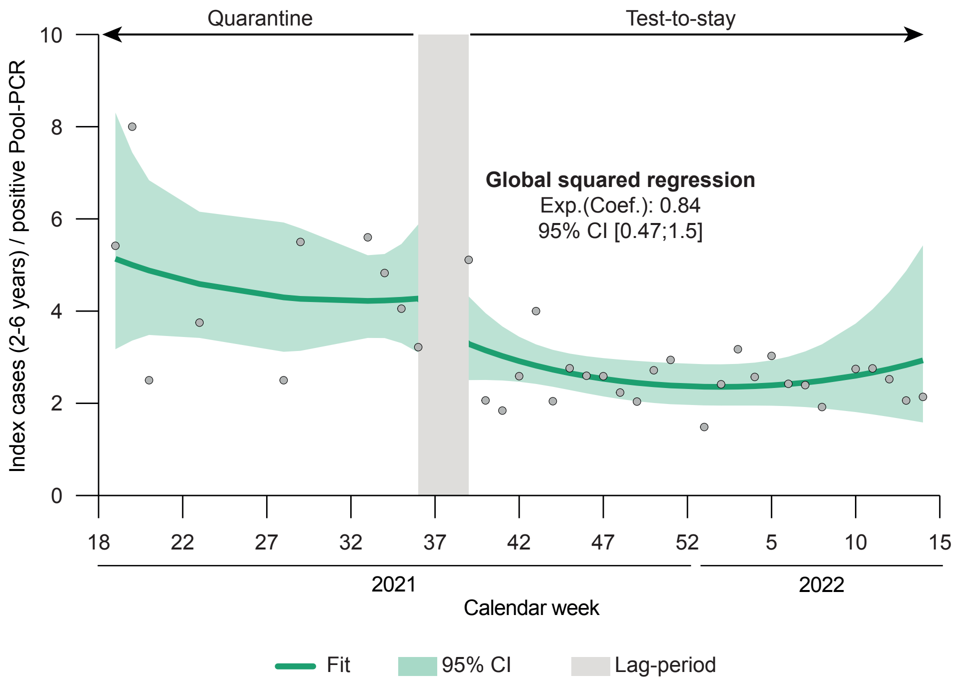


**Supplemental Figure 10: RDiT analysis using global squared regression instead of global linear regression**
The outcome is fit assuming a gamma distribution. The exponentiated coefficient is interpreted on a multiplicative scale.

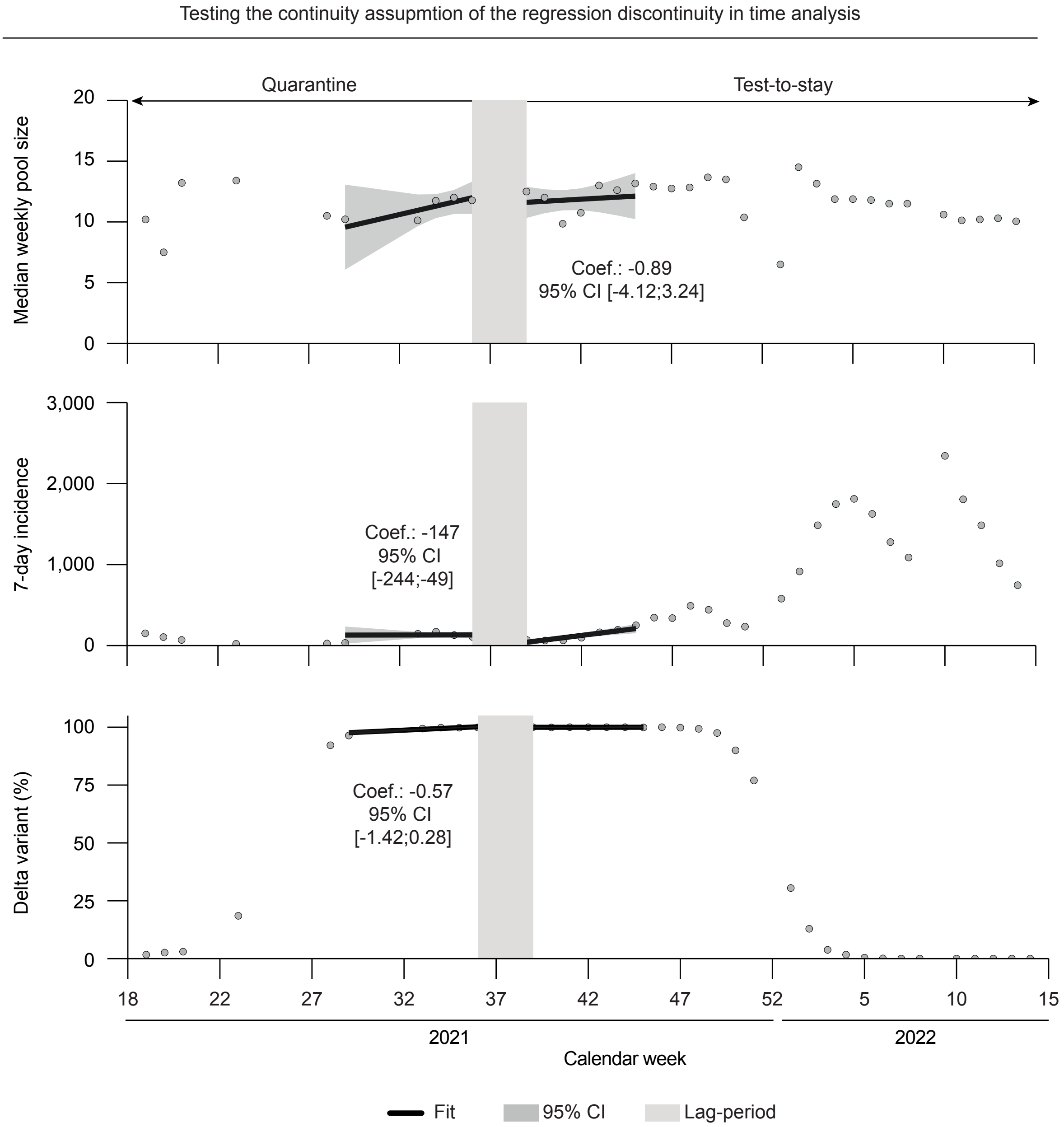


**Supplemental Figure 11: Testing continuity of baseline covariates**

Local linear regressions for pool size (top), 7-day incidence (middle), and share of the Delta variant (bottom) to test the RDiT continuity assumption for baseline covariates. As outcomes are fit assuming a normal distribution, the coefficients are interpreted on an additive scale.

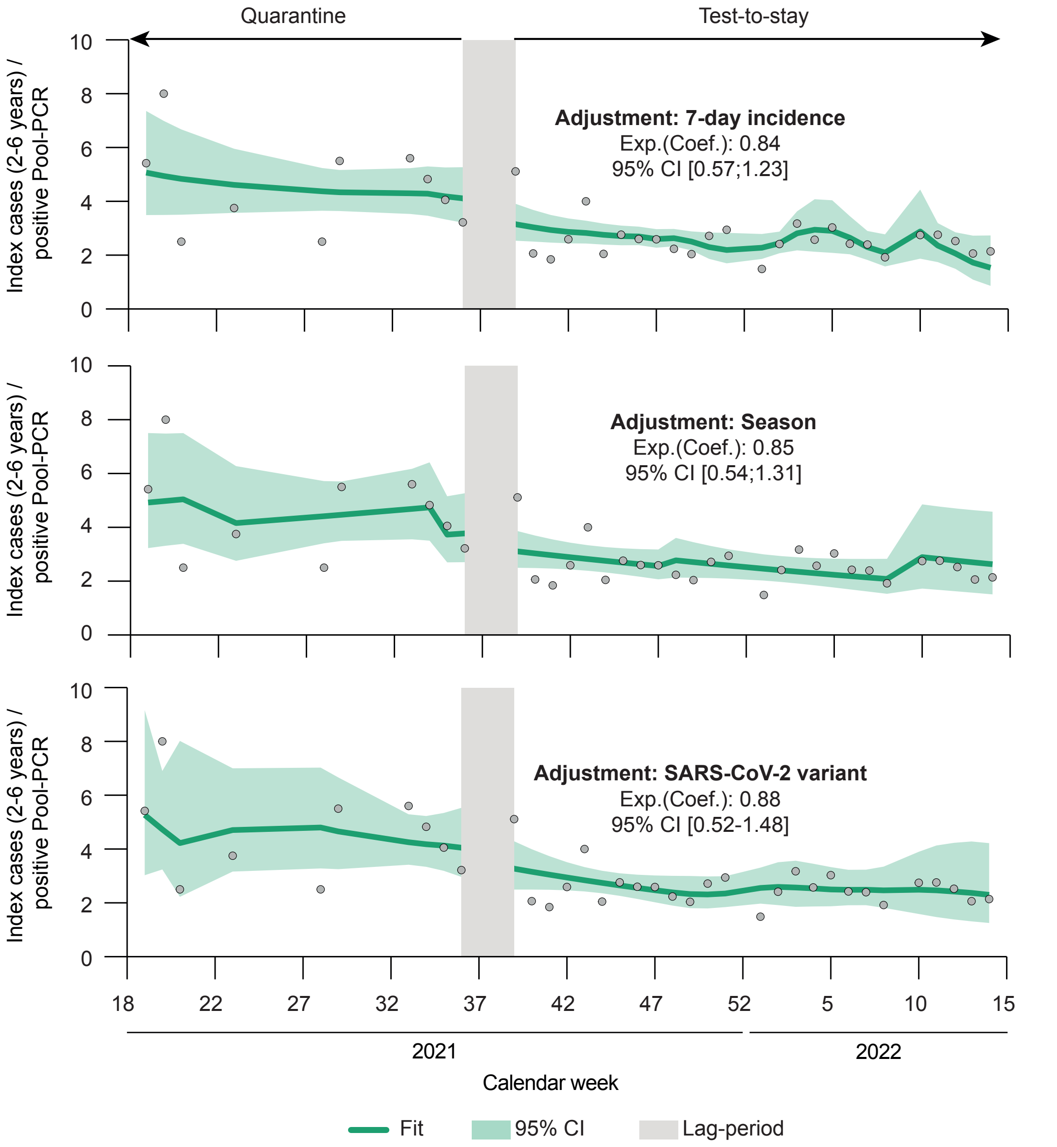


**Supplemental Figure 12: Sensitivity analyses 1**

Global linear regression including 7-day incidence (continous), season (spring, summer, autumn, winter as dummy variables), and SARS-CoV-2 variant (continous) as covariates in the model specification. The outcome is fit assuming a gamma distribution.The exponentiated coefficients are interpreted on a multiplicative scale.

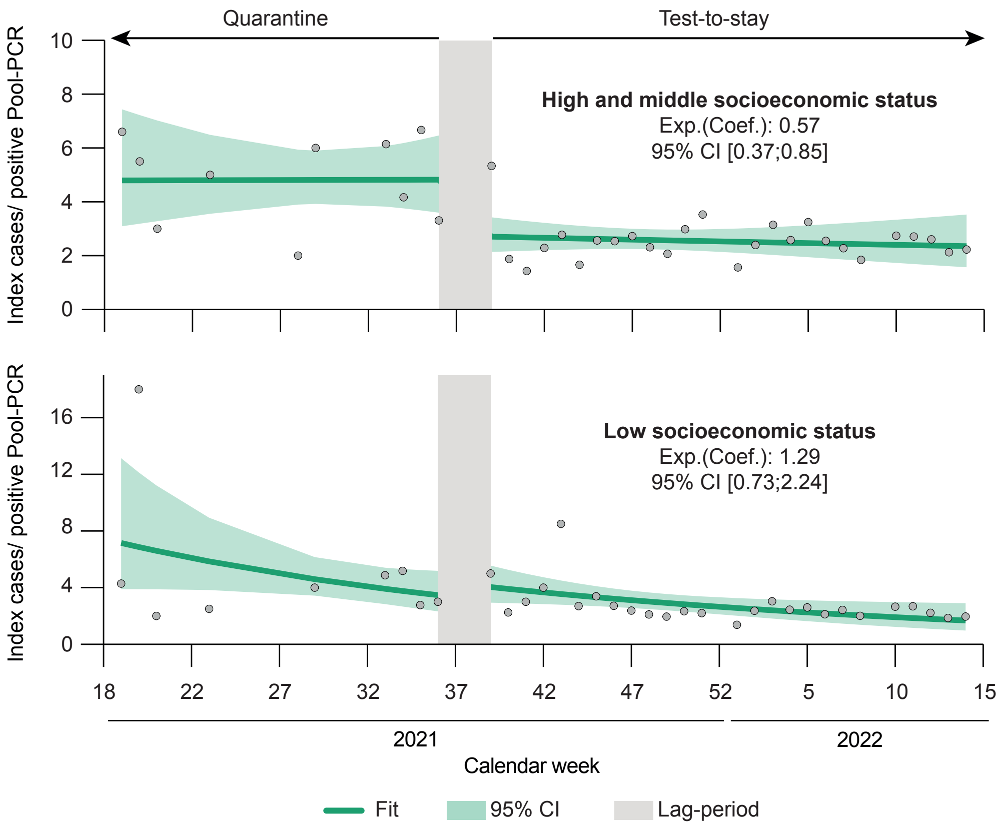


**Supplemental Figure 13: Sensitivity analyses 2**

Global gamma regressions stratified by socioeconomic status of the city districts (high/middle, top and low, bottom). The outcome is fit assuming a gamma distribution.The exponentiated coefficients are interpreted on a multiplicative scale.
